# SARS-CoV-2 DNA Vaccine INO-4800 Induces Durable Immune Responses Capable of Being Boosted in a Phase 1 Open-Label Trial

**DOI:** 10.1101/2021.10.06.21264584

**Authors:** Kimberly A. Kraynyak, Elliott Blackwood, Joseph Agnes, Pablo Tebas, Mary Giffear, Dinah Amante, Emma L. Reuschel, Mansi Purwar, Aaron Christensen-Quick, Neiman Liu, Viviane M. Andrade, Malissa C. Diehl, Snehal Wani, Martyna Lupicka, Albert Sylvester, Matthew P. Morrow, Patrick Pezzoli, Trevor McMullan, Abhijeet J. Kulkarni, Faraz I. Zaidi, Drew Frase, Kevin Liaw, Trevor R.F. Smith, Stephanie J. Ramos, John Ervin, Mark Adams, Jessica Lee, Michael Dallas, Ami Shah Brown, Jacqueline E. Shea, J. Joseph Kim, David B. Weiner, Kate E. Broderick, Laurent M. Humeau, Jean D. Boyer, Mammen P. Mammen

## Abstract

**Background:** Additional SARS-CoV-2 vaccines that are safe and effective as primary vaccines and boosters remain urgently needed to combat the COVID-19 pandemic. We describe the safety and durability of the immune responses following two primary doses and a homologous booster dose of an investigational DNA vaccine (INO-4800) targeting the full-length spike antigen.

**Methods:** Three dosage strengths of INO-4800 (0.5 mg, 1.0 mg, and 2.0 mg) were evaluated in 120 age-stratified healthy adults. Intradermal injection of INO-4800 followed by electroporation at 0 and 4 weeks preceded an optional booster 6-10.5 months after the second dose.

**Results:** INO-4800 appeared well tolerated, with no treatment-related serious adverse events. Most adverse events were mild and did not increase in frequency with age and subsequent dosing. A durable antibody response was observed 6 months following the second dose; a homologous booster dose significantly increased immune responses. Cytokine producing T cells and activated CD8+ T cells with lytic potential were significantly increased in the 2.0 mg dose group.

**Conclusion:** INO-4800 was well tolerated in a 2-dose primary series and as a homologous booster in all adults, including the elderly. These results support further development of INO-4800 for use as a primary vaccine and as a booster.

Trial Registration: ClinicalTrials.gov NCT04336410

**Summary:** Two-milligram dose of INO-4800, a DNA-based vaccine encoding the SARS-CoV-2 spike protein, appears safe and well-tolerated and elicits humoral and cell-mediated immunity persisting to 6 months after a second dose. A third dose 6-10.5 months later significantly boosts immune responses.

## Introduction

Despite aggressive vaccination campaigns, most of the world’s population remains unvaccinated and susceptible to COVID-19, the disease caused by SARS-CoV-2[1]. The urgent need remains for additional safe and effective vaccines that are affordable, scalable, and can be distributed to countries where the infrastructure may not be supportive of ultra-cold chain transport and storage.

Attachment of SARS-CoV-2 to host cells is mediated by binding of the viral spike (S) protein to angiotensin converting enzyme 2 (ACE2) receptors on host cells[2]. Humoral responses against the spike protein prevent the virus from accessing host cells[3], and this strategy has led to the development of several vaccines targeting SARS-CoV-2 (reviewed by[4, 5]).

INO-4800 is an investigational optimized DNA vaccine, encoding the SARS-CoV-2 S protein[6], injected intradermally followed by in vivo electroporation[7]. This approach potentially offers several advantages, including induction of humoral and cellular immunity, favorable tolerability and thermal stability profiles, and ease of manufacture[8, 9]. Plasmid DNA-based products in development by this sponsor have been shown to be stable at 2-8°C for 3-5 years, at room temperature (25°C) for least 1 year, and at 37°C for 1 month (unpublished data), and is in line with earlier reports on the stability of pharmaceutical grade plasmid DNA[10].

Preclinical studies have shown INO-4800 to be immunogenic[6], with durable cellular and neutralizing antibody responses[11]. INO-4800 provided protection against viral challenge in non-human primates with no evidence of vaccine-enhanced disease[12], and elicited neutralizing antibodies reactive against multiple variants of concern (VOCs)[13].

The preliminary safety and immunogenicity of INO-4800 in both Phase 1 and Phase 2 clinical studies have been previously reported[14, 15]. The earlier analysis[14] demonstrated that two doses of INO-4800 administered one month apart were well tolerated in 38 healthy participants 18-50 years of age and induced neutralizing antibodies and/or T-cells. Here we describe the durability of that response at 6 months following the second dose, as well as the safety and immunogenicity of the 2-dose regimen in older and elderly participants, including following a subsequent homologous booster dose.

## Methods

### Trial Design and Participants

This Phase 1, open-label, multi-center trial (NCT04336410) evaluated the safety, tolerability, and immunogenicity of INO-4800 injected intradermally (ID) followed by electroporation (EP). A total of 120 healthy participants without a known history of COVID-19 were assigned to receive a 0.5mg, 1.0mg, or 2.0mg dose of INO-4800 in a 2-dose regimen (weeks 0 and 4) and a subsequent optional booster dose no earlier than 8 weeks after dose 2. An equal number of participants were enrolled in each dose group (n=40) and further stratified by age groups [18-50 years of age; n=20, 51-64 years of age; n=10, and ≥65 years of age; n=10].

The trial was approved by the institutional review board of each clinical site, all participants provided written informed consent prior to enrollment. The trial was conducted under current Good Clinical Practices (GCP).

### DNA Vaccine INO-4800

INO-4800 was previously described[6, 14] and encodes the full-length sequence of the SARS-CoV-2 spike glycoprotein derived from the Wuhan strain based on an optimized synthetic sequence created using a proprietary algorithm. The final vaccine drug product, manufactured under Good Manufacturing Practices, was formulated at 10mg/mL in saline sodium citrate buffer.

INO-4800 is injected ID followed by EP using the CELLECTRA® 2000 device that generates a controlled electric field at the injection site to enhance the cellular uptake and expression of the DNA plasmid as previously described[16, 17]. The device delivers a total of four electrical pulses per EP, each of 52 msec in duration, at current of 0.2 Amp and voltage of 40-200 per pulse.

### Endpoints

Primary safety endpoints included incidence of adverse events (AEs) using the “Toxicity Grading Scale for Healthy Adult and Adolescent Volunteers Enrolled in Preventive Vaccine Clinical Trial” including frequency and severity of injection site reactions. Primary immunological endpoints included the measurement of SARS-CoV-2 Spike glycoprotein antigen-specific binding antibodies as well as the measurement of antigen-specific cellular immune responses by IFN-γ, ELISPOT and flow cytometry assays. Endpoints reflected in this publication are inclusive of 6 months after second dose (non-boosted participants) and, when applicable, 2 weeks after booster dose.

### Trial Procedures

Vaccine was administered in 0.1 ml ID injections in the deltoid followed by EP at the injection site. At each dosing visit, either a single injection for 0.5mg and 1.0mg dose groups or two injections for 2.0mg dose group were given, one in each deltoid.

Forty participants 18-50 years were enrolled sequentially into 1.0mg and 2.0mg dose groups with a safety run-in period[14]. The trial design was expanded to include older participants in all dosing groups (including a 0.5mg dose level). Upon favorable safety assessment review by an independent Data Safety Monitoring Board (DSMB) of Week 1 data for 0.5mg dose group participants aged 51-64 years and ≥ 65 years, enrollment of the corresponding age strata in the 1.0mg and subsequently 2.0mg dose groups was initiated.

Participants were assessed for safety (complete blood count, serum chemistry, and urinalysis), including local and systemic AEs, at screening, Week 0 (Dose 1), next day phone call, and Weeks 1, 4 (Dose 2), 6, 8, 12, 28, 40 and 52. Blood immunology collections occurred at all clinic visits except Week 1. After the Week 12 visit, participants who consented to the optional booster dose were transitioned to an extended schedule of events to include the booster dose (Dose 3) and subsequent visits for safety at 2, 12, 24, 36, and 48 weeks following the booster dose with blood immunology collections at all clinic visits except 36 weeks.

The DSMB reviewed laboratory and AE data for the participants up to 24 weeks after the second dose (non-boosted) and 2 weeks after booster dose.

### Protocol Eligibility

Key inclusion criteria included: healthy adults aged at least 18 years; and Body Mass Index of 18-30kg/m^2^ at screening. Key exclusion criteria included: individuals in a current occupation with high risk of exposure to SARS-CoV-2; previous known exposure to SARS-CoV-2 or receipt of an investigational product for the prevention or treatment of COVID-19; autoimmune or immunosuppression as a result of underlying illness or treatment; hypersensitivity or severe allergic reactions to vaccines or drugs; and medical conditions that increased risk for severe COVID-19.

### Immunogenicity Assessment Methods

Samples were collected at timepoints described above with screening and pre-dose 1 samples considered baseline. Peripheral blood mononuclear cells (PBMCs) were collected as previously described[14]. After isolation, PBMCs were stored in the vapor phase of a liquid nitrogen freezer until analysis, while serum samples were stored at −80°C. Eight participants were excluded from the immunogenicity analyses due to a positive ELISA titer to the SARS-CoV-2 nucleoprotein, suggesting SARS-CoV-2 infection

### SARS-CoV-2 Pseudovirus Neutralization Assay

Serum samples were measured using a pseudovirus neutralization assay as described previously[15]. Data was reported as ID_50_, which is the reciprocal serum dilution resulting in 50% inhibition of infectivity by comparison to control wells with no serum samples added. Supplementary methods show additional information.

### SARS-CoV-2 Spike Enzyme-Linked Immunosorbent Assay (ELISA)

Binding antibodies to SARS-CoV-2 spike protein were measured by ELISA as described previously[15]. SARS-CoV-2 spike antibody concentrations were determined by interpolation from a dilution curve of SARS-CoV-2 convalescent plasma with an assigned concentration of 20,000 Units/mL. Supplementary methods show additional information.

### SARS-CoV-2 Spike ELISpot Assay Description

The SARS-CoV-2 spike antigen-specific IFN-γ T-cell response was measured as described previously[14]. Values were reported as the mean spot-forming units per million PBMCs across three triplicate wells after background subtraction using DMSO-only negative control wells. Supplementary methods show additional information.

### INO-4800 SARS-CoV-2 Spike Flow Cytometry Assays

PBMCs were also assessed in Intracellular Cytokine Staining (ICS) and Lytic Granule Loading (LGL) assays. The ICS assay was performed as previously described[14]. The LGL assay was also performed as reported previously[18] following stimulation with overlapping peptides to the full-length spike protein to measure CD8+T cell activation and capacity to produce lytic proteins.

### Statistical Analysis

No formal power analysis was applicable to this trial. Descriptive statistics were used to summarize the safety endpoints based on the safety population: proportions of participants with AEs, through 6 months following dose 2 (non-boosted participants) or 2 weeks following booster dose. The safety population included all participants who received at least one dose of INO-4800 and were grouped by age and the dose of INO-4800. Post-hoc within subject analyses of post-vaccination minus pre-vaccination paired differences in SARS-CoV-2 neutralization and ELISA spike responses (on the natural log-scale, with a paired t-test), ELISpot responses (with Wilcoxon signed-rank tests), and flow assay responses (with Wilcoxon signed-rank tests) were performed.

## Results

### Trial Population Demographics

Between 06 April 2020 and 07 July 2020, 154 participants were screened and 120 enrolled into the trial (**Figure 1**). The median age was 50.5 years (range 18 to 86 years). Participants were 57.5% female (69/120) and 42.5% male (51/120) (**Table 1**). Most participants were white (94.2%, 113/120).

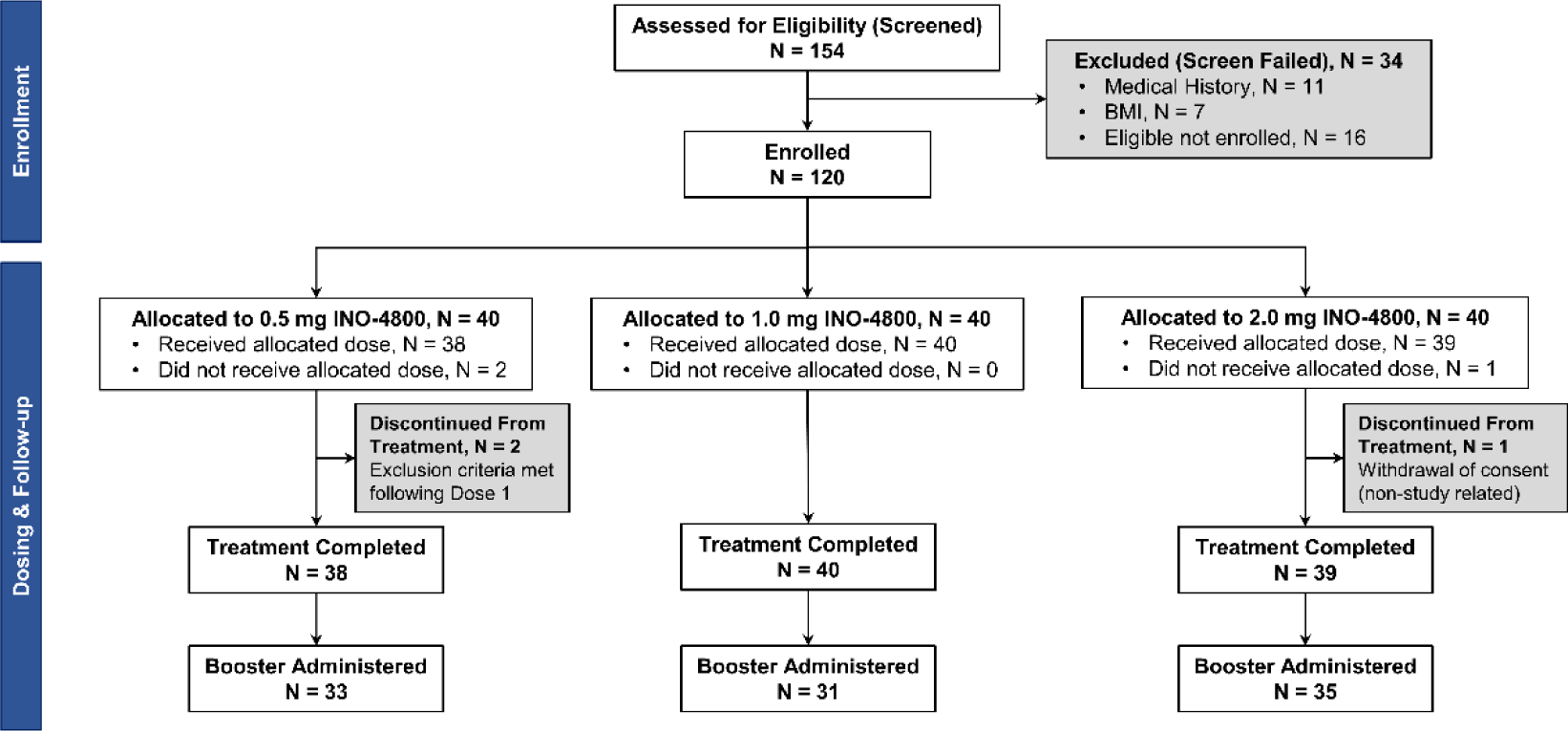

**Figure 2.**
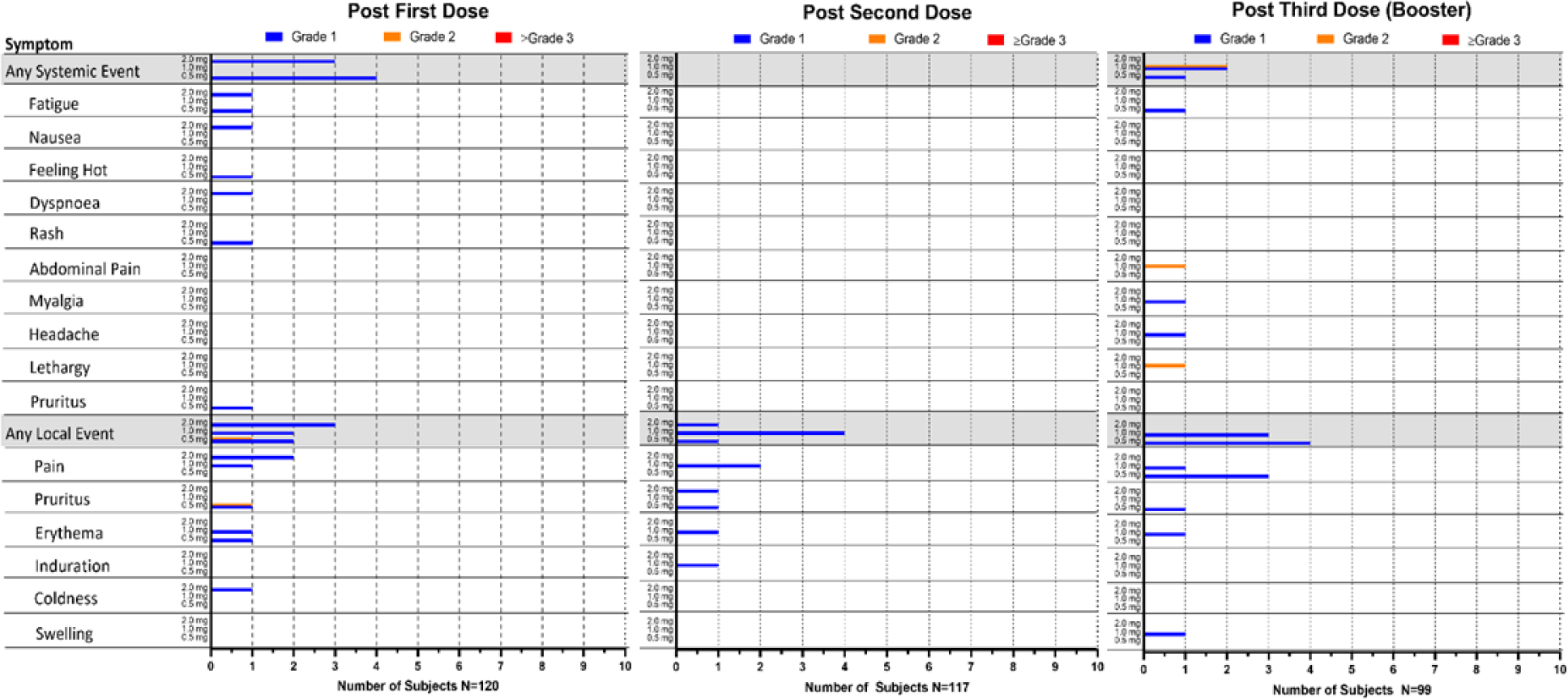
Related systemic and local adverse events. Post First Dose, N=120 (N=40 in each dose group), Post Second Dose, N=117 (N=38 in the 0.5 mg dose group, N=40 in the 1.0 mg dose group and N=39 in the 2.0 mg dose group, and Post Third Dose, N=99 (N=33 in the 0.5 mg dose group, N=31 in the 1.0mg dose group and N=35 in the 2.0 mg dose group)

**Table 1.** Participant Demographics

| Variable | Statistic | 0.5 mg<br>(N=40) | 1mg<br>(N=40) | 2mg<br>(N=40) | Total<br>(N=120) |
| --- | --- | --- | --- | --- | --- |
| **Sex** |  |  |  |  |  |
| **Male** | n (%) | 18 (45.0) | 17 (42.5) | 16 (40.0) | 51 (42.5) |
| **Female** | n (%) | 22 (55.0) | 23 (57.5) | 24 (60.0) | 69 (57.5) |
| **Race** |  |  |  |  |  |
| **White** | n (%) | 40 (100) | 38 (95.0) | 35 (87.5) | 113 (94.2) |
| **Black or African**<br>**American** | n (%) | 0 | 1 (2.5) | 1 (2.5) | 2 (1.7) |
| **Asian** | n (%) | 0 | 1 (2.5) | 4 (10.0) | 5 (4.2) |
| **Ethnicity** |  |  |  |  |  |
| **Hispanic or Latino** | n (%) | 3 (7.5) | 0 | 0 | 3 (2.5) |
| **Not Hispanic or**<br>**Latino** | n (%) | 35 (87.5) | 40 (100) | 40 (100) | 115 (95.8) |
| **Not Reported** | n (%) | 2 (5.0) | 0 | 0 | 2 (1.7) |
| **Age (years)** | N | 40 | 40 | 40 | 120 |
|  | Mean (SD) | 50.7 (15.30) | 49.2<br>(16.75) | 50.7<br>(17.90) | 50.2 (16.56) |
|  | Median | 52.5 | 51.0 | 50.5 | 50.5 |
|  | Min, Max | 23, 76 | 18, 73 | 19, 86 | 18, 86 |

### Vaccine Safety and Tolerability

A total of 117 of 120 (97.5%) participants received both doses. One participant in the 2.0 mg group discontinued trial participation prior to receiving the second dose solely due to lack of transportation to the clinical site. Two participants in the 0.5 mg group did not receive the second dose due to exclusionary eligibility criteria (hypertension) having been determined following Dose 1; (**Figure 1**).

Ninety-nine of 120 (82.5%) participants consented to and received the booster dose, approximately 6 to 10.5 months following the second dose. Reasons for not receiving booster dose included receipt of another SARS-CoV-2 vaccine (available under Emergency Use Authorization), new medical condition precluding participation (having had COVID-19, pregnancy or hypertension), or loss to follow-up.

A total of 34 treatment-related local and systemic AEs were reported by 18 participants. Thirty-one AEs were Grade 1 (mild) in severity and comprised mostly injection site reactions. Three treatment-related Grade 2 (moderate) AEs were reported as lethargy, abdominal pain, and injection site pruritus. There were no febrile reactions reported. No participants discontinued due to AEs. No treatment-related SAEs were reported. There were no abnormal laboratory values that were deemed treatment-related and clinically significant by the Investigators. There was no increase in the number of participants who experienced related AEs in the 2.0 mg group (12.5%, 5/40), compared to that in the 1.0 mg group (15%, 6/40) or the 0.5 mg group (17.5%, 7/40). In addition, there was no appreciable increase in the frequency of AEs with the second or booster doses when compared to the first dose (**Figure 2**). A decrease in frequency of treatment-related AEs in the older and elderly age cohorts was observed when compared to the younger age group (**Supplementary Table 4**).

### INO-4800 induces durable humoral immune responses capable of being boosted

Induction of antibodies against SARS-CoV-2 following vaccination with INO-4800 was measured from sera. The functionality of antibodies was assessed using a pseudovirus neutralization assay. All dose groups induced neutralizing antibodies that peaked two weeks post second dose (GMTs-14.9, 19.1, 54.1 in the 0.5 mg, 1.0 mg and 2.0 mg dose groups, respectively) (**Figure 3A, left panel, Supplementary Table 1**). These increased responses were statistically significant over baseline in the 2.0 mg dose group for each time point through study week 28, approximately 6 months after dose 2 (**Figure 3A, table**). Following administration of a booster dose, statistically significant increases over pre-boost titers were observed in all dose groups (GMTs-58.7, 76.1, 100 in the 0.5 mg, 1.0 mg and 2.0 mg dose groups, respectively; all P<0.001) (**Figure 3A, right panel, Supplementary Table 1**). The 2.0 mg dose group had a 12.8 (95%CI 6.3, 26.0) geometric fold rise (GMFR) over pre-boost titers, the highest of any dose group. Neutralization titers by participant age are shown in **Supplementary Figure 1A;** GMTs were numerically lower in the older age groups but statistically significantly higher than baseline at week 6 in the 2.0 mg dose group. Plasma samples from convalescent samples had a GMT of 922 and ranged from 10 to 13,249 **(Supplementary Figure 2A)**.

**Figure 3.**
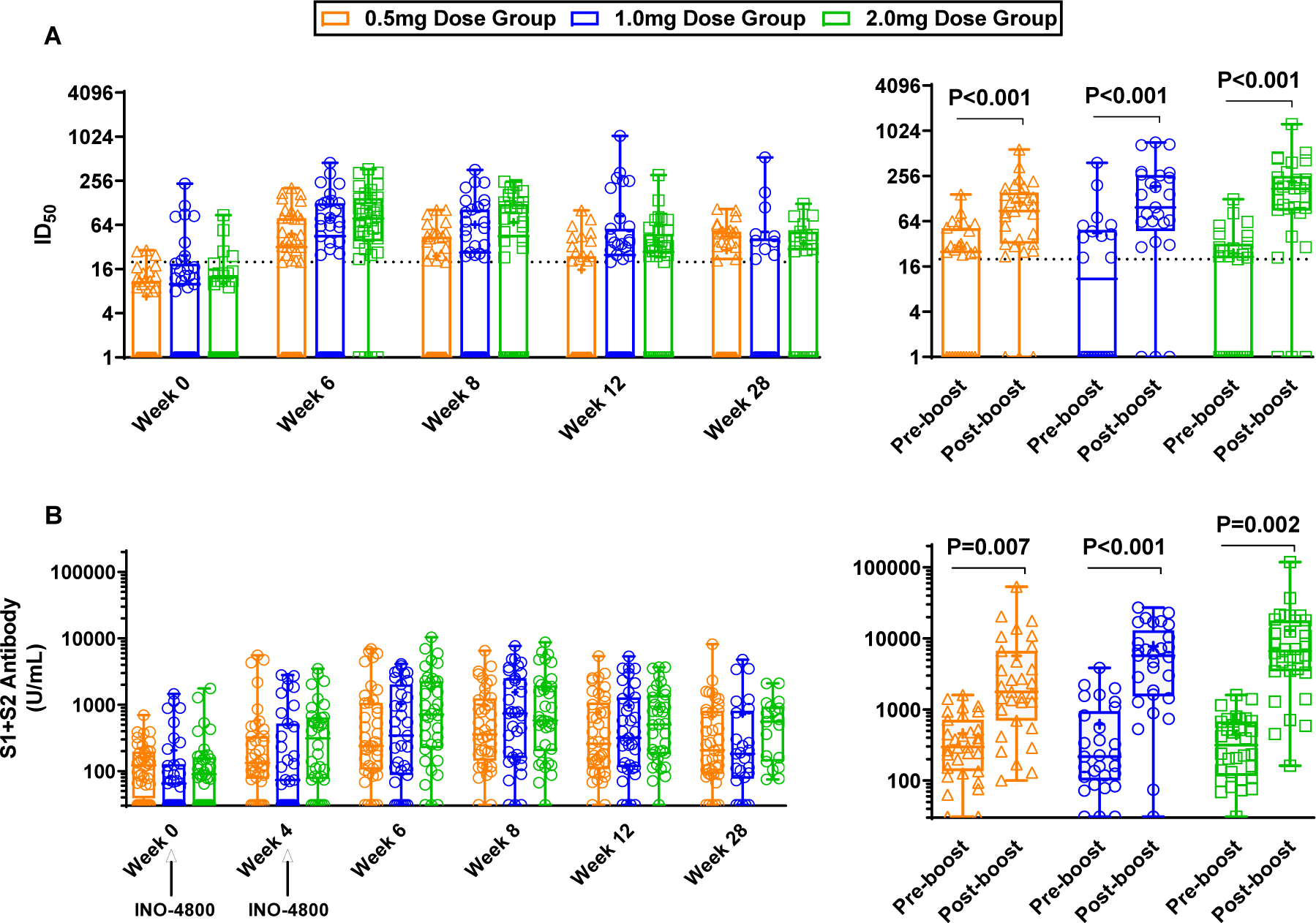
INO-4800 induces antibodies to SARS-CoV-2. A) Functional antibodies were assessed using a pseudovirus neutralization assay. The inhibition dilution where 50% neutralization occurs (ID_50_) is plotted. The dotted line represents the lowest dilution tested in the assay (1:20). The left panel includes n=40 participants in the 0.5 mg dose group, n=35 participants in the 1.0 mg dose group and n=36 participants in the 2.0 mg dose group. The right panel includes n=33, n=26 and n=31 participants in the 0.5 mg, 1.0 mg, and 2.0 mg dose groups, respectively. B) Binding antibody concentrations to the Spike trimer were measured using ELISA. The left panel includes n=40 participants in the 0.5 mg dose group, n=35 participants in the 1.0 mg dose group and n=36 participants in the 2.0 mg dose group. The right panel includes n=31, n=29 and n=32 participants in the 0.5 mg, 1.0 mg, and 2.0 mg dose groups, respectively. Open symbols represent individual participants, the box extends from the 25^th^ to the 75^th^ percentile, line inside the box is the median, and the whiskers extend from the minimum to maximum values. The mean is denoted with a “+” sign. Paired t test was used to assess significance versus baseline. The dose groups are represented by orange triangles (0.5 mg), blue circles (1.0 mg) and green squares (2.0 mg).

Antibodies to the spike trimer protein were measured in a binding ELISA. All three groups exhibited binding antibodies that peaked four weeks following dose 2 (Geometric Mean Titers, GMTs-428.5, 595.9, 678.0 in the 0.5 mg, 1.0 mg and 2.0 mg dose groups, respectively) (**Figure 3B, left panel, Supplementary Table 2**). Increases over baseline were observed in all participants who received the 2.0 mg dose, but not in all participants in the other groups, and GMTs were statistically significantly higher than baseline 6 months following dose 2 (GMTs-250.1, 215.3, 407.2 in the 0.5 mg, 1.0 mg and 2.0 mg dose groups, respectively; all P<0.026).

Following administration of a booster dose, statistically significant increases over pre-boost titers were observed in all dose groups (GMTs-1963.8, 3685, 5953 in the 0.5 mg, 1.0 mg and 2.0 mg dose groups, respectively; all P≤0.007) (**Figure 3B, right panel, Supplementary Table 2**). The 2.0 mg dose group had a 20.8 (95%CI 13.9, 31.2) GMFR over pre-boost titers which was the highest of any dose group. ELISA binding titers by participant age are shown in **Supplementary Figure 1B.** Plasma samples from convalescent samples had a GMT of 19,444 and ranged from 330 to 247,200 **(Supplementary Figure 2B)**.

### INO-4800 induces cellular immune responses capable of being boosted

Interferon-gamma (IFNγ) ELISpot was performed on PBMCs. Increases in spot forming units (SFU) per million PBMCs over baseline are shown in **Figure 4A, left panel**. Magnitudes of IFNγ peaked at week 6 for the 0.5 mg and 2.0 mg dose groups (median 19.4 and 43.3, respectively) and at week 8 for the 1.0 mg dose group (median 17.8). Six months following dose 2, magnitudes remained high in the 2.0 mg dose group (median 19.6). Of note, magnitudes in the 1.0 mg and 2.0 mg dose groups were statistically significantly increased following the booster dose (P=0.018 and P=0.008, respectively) (**Figure 4A, right panel**). The 2.0 mg dose group had a difference in medians of 10 following the booster, resulting in the highest post-boost increase of any dose group. ELISpot responses by participant age are shown in **Supplementary Figure 3A.**

**Figure 4.**
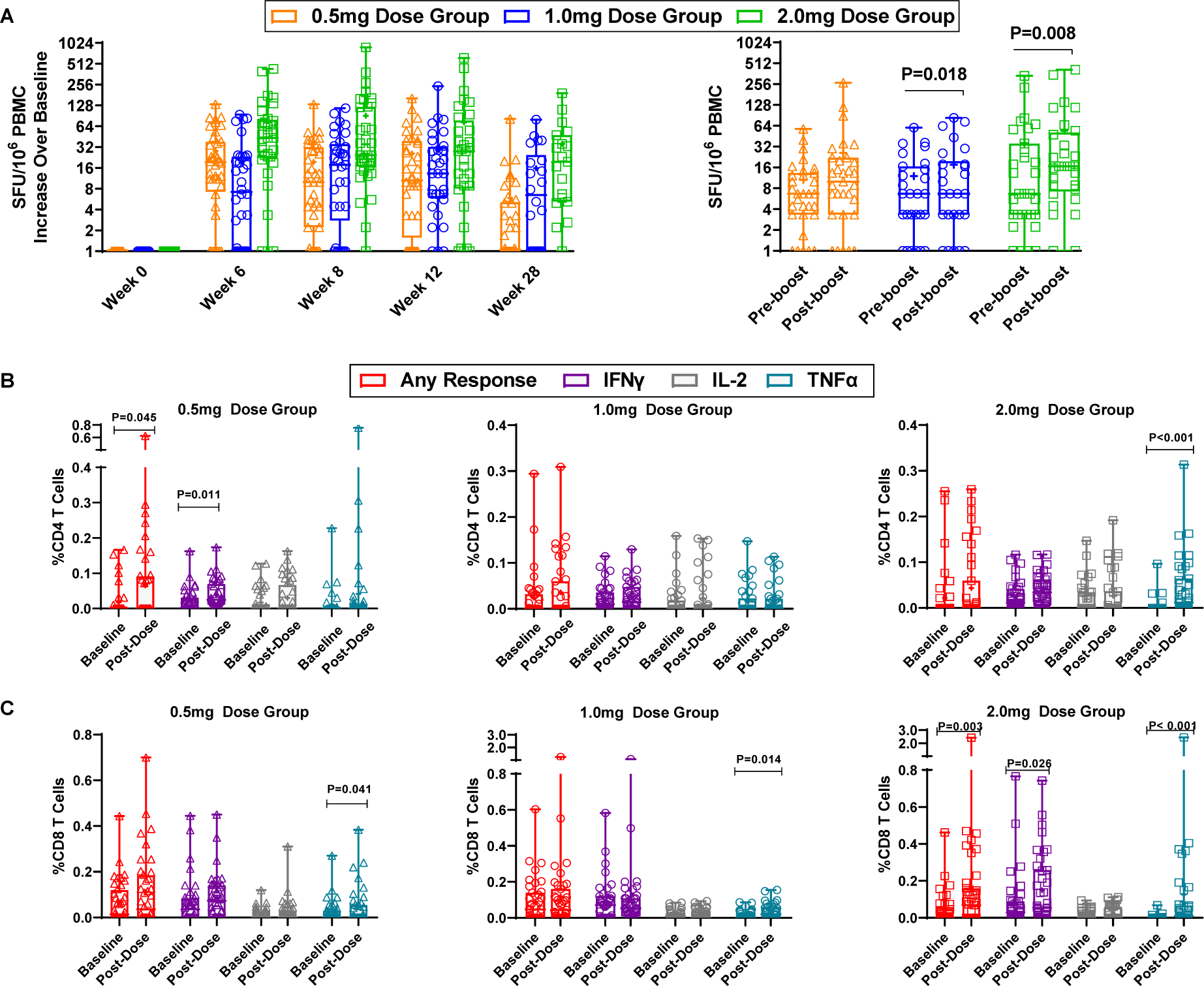
INO-4800 induces cellular responses to SARS-CoV-2 Spike. A) Longitudinal increases in spike antigen specific spot forming units per 10^6^ PBMCs over baseline in the IFN-g ELISpot are plotted. The left panel includes n=40 participants in the 0.5 mg dose group, n=35 participants in the 1.0 mg dose group and n=36 participants in the 2.0 mg dose group. The right panel includes n=31, n=30 and n=34 participants in the 0.5 mg, 1.0 mg, and 2.0 mg dose groups, respectively. B-C) Intracellular cytokine staining for IFN-g (purple) IL-2 (gray), TNF-a (blue) or any of the three cytokines (red) are plotted from samples collected at baseline or post-dose 2. The graphs include n=40 participants in the 0.5 mg dose group and n=39 participants in the 1.0 mg and 2.0 mg dose groups. Open symbols represent individual participants, the box extends from the 25^th^ to the 75^th^ percentile, line inside the box is the median, and the whiskers extend from the minimum to maximum values. The mean is denoted with a “+” sign. Wilcoxon signed-rank was used to assess significance versus baseline. The dose groups are represented by triangles (0.5 mg), circles (1.0 mg) and squares (2.0 mg).

### INO-4800 induces cytokine producing T cells and activated CD8+T cells with lytic potential

The contribution of SARS-CoV-2 specific CD4^+^ and CD8^+^ T cells was assessed by intracellular cytokine staining (ICS) on participants following 2 doses, **Figure 4B-C**. The median frequency of CD4+T cells producing IFNγ increased following vaccination in all three dose groups of INO-4800, and the frequency of CD4+T cells producing TNFα was statistically significantly increased in the 2.0 mg dose group (P<0.001) (**Figure 4B**). The frequency of CD8+T cells producing TNFα was statistically significantly increased following vaccination in all three dose groups of INO-4800 (All P≤0.041) (**Figure 4C**). The 2.0 mg dose group had the highest difference in medians for CD8+T cells producing any response, IFNγ and TNFα (0.066, 0.026, and 0.011 respectively). Responses by participant age are shown in **Supplementary Figure 3B-C.**

SARS-CoV-2 specific CD8+T cells were also characterized on a subset of participants with remaining sample following 3 doses by a flow cytometry assay that included activation markers CD69 and CD137. The median frequency of CD8+CD69+CD137+ cells increased following immunization with 2.0 mg of INO-4800, with a difference in the medians of 0.072 **(Figure 5A, left panel)**. Further characterization of these activated cells, including the co-expression of proteins utilized in cytolytic killing (granzyme A, granzyme B, perforin or granulysin) revealed a statistically significant increase in both the 1.0 mg (P=0.008) and 2.0 mg (P=0.003) dose groups **(Figure 5A middle and right panels).** The 2.0 mg dose group had a difference in medians of 0.085 in the CD69+CD137+ population co-expressing perforin and granzymes A and B and 0.054 in the population co-expressing granulysin. CD8+T cells expressing the activation marker CD38 and proliferation marker Ki67 were also assessed **(Figure 5B and C, respectively).** The frequency of SARS-CoV-2 specific CD38+CD8+T cells statistically significantly increased following 2.0 mg of INO-4800 (P=0.016), with a difference in medians of 1.45 (**Figure 5B, left panel**). CD38+CD8+T cells with lytic potential (**Figure 5B middle and right panels**) statistically significantly increased following 2.0 mg of INO-4800 (P<0.001). Following immunization with 2.0 mg of INO-4800, the mean frequency of activated CD8+T cells expressing granzymes A and B and perforin was 1.7% with a difference in medians of 0.710 and those expressing granulysin was 1.8% with a difference in medians of 0.433 (**Figure 5B middle and right panels)**. Statistically significant increases in the frequency of these CTL phenotypes were also observed in the 1.0 mg dose group (P≤0.012) (**Figure 5B middle and right panels**). The 2.0 mg dose group had the highest frequencies of CD8+T cells expressing Ki67 with a difference in medians of 0.367 and Ki67 with cytolytic proteins: 0.296 (GrzA+GrzB+Prf+) and 0.230 (Gnly+). All three Ki67+ populations were statistically significantly increased in the 2.0 mg dose group (P≤0.001; **Figure 5C)**. The 2.0 mg dose group consistently showed the highest median responses across all phenotypes assessed compared to the other groups.

**Figure 5.**
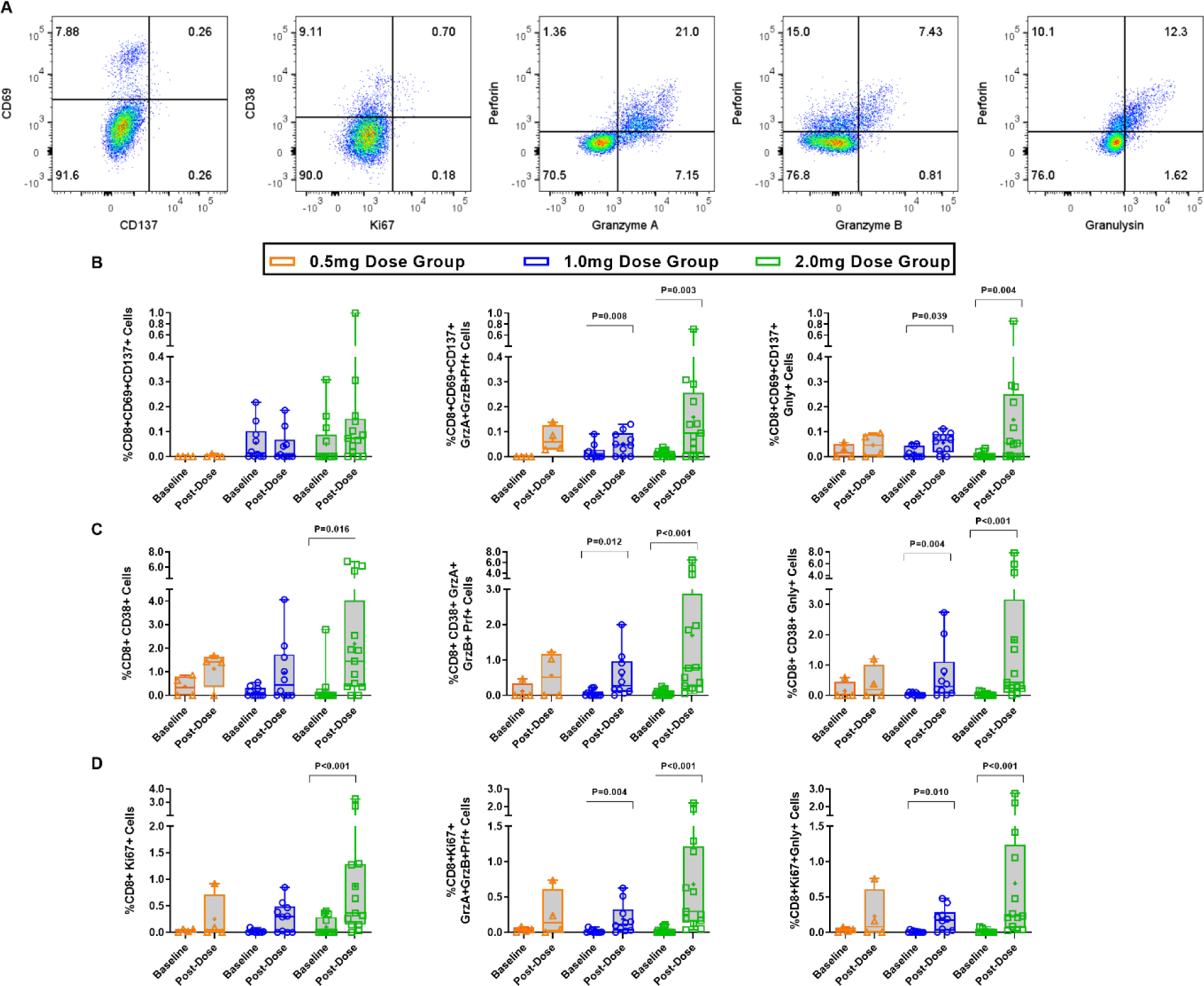
INO-4800 induces spike specific activated CD8+T cells with lytic potential. A lytic granule loading flow cytometry assay was used to further characterize CD8+T cells and an example gating strategy is shown in (A). The expression of the activation markers CD69 and CD137 (B), CD38 (C), and the proliferation marker Ki67 (D) from samples collected at baseline or post-dose 2. The expression of proteins found in lytic granules: granzymes A (GrzA) and B (GrzB), perforin (Prf) and granulysin (Gnly) were assessed together with activation/proliferation subset. The graphs include n=4 participants in the 0.5 mg dose group and n=10 participants in the 1.0 mg dose group and n=13 in the 2.0 mg dose group. Open symbols represent individual participants, the box extends from the 25^th^ to the 75^th^ percentile, line inside the box is the median, and the whiskers extend from the minimum to maximum values. The mean is denoted with a “+” sign. Wilcoxon signed-rank was used to assess significance versus baseline. The dose groups are represented by orange triangles (0.5 mg), blue circles (1.0 mg) and green squares (2.0 mg).

## Discussion

This report provides results for the expansion of a Phase 1 trial to include older and elderly participants and an optional booster dose with the aim to evaluate the safety, tolerability, and immunogenicity of INO-4800, a SARS-CoV-2 vaccine encoding the S protein[14], including the immune responses 6 months following dose 2 and 2 weeks following the optional booster dose.

INO-4800 appeared to be well-tolerated at all three dose levels, with no treatment-related serious adverse events (SAEs) reported. Most AEs were mild in severity and did not increase in frequency with age and subsequent dosing. These results are consistent with the severity of AEs and lack of treatment-related SAEs observed in the U.S. Phase 2 trial comparing the 1.0 mg and 2.0 mg doses of INO-4800 in approximately 400 subjects[15] and those studies conducted outside the U.S. by Inovio collaborators (International Vaccine Institute, Advaccine – manuscripts in preparation). The lower frequency of treatment-related AEs reported by older and elderly participants in our study is consistent with findings of other studies evaluating SARS-CoV-2 vaccines[19, 20]. Weaker inflammatory reactions consequent to immunosenescence may explain the lower incidence of AEs among elderly participants[21, 22].

Induction of both humoral and cellular responses were observed across all three dose groups, inclusive of binding and neutralizing antibodies and cytokine producing T cells as well as exhibiting lytic potential. Immunization with the 2.0 mg dose resulted in the highest GMTs of neutralizing and binding antibodies as well as the highest magnitudes of IFNγ production to SARS-CoV-2 of any dose in all age groups tested, and the increases in antibody levels were statistically significant above baseline at 6 months following dose 2. Importantly, increases in both humoral and cellular immune responses were statistically significant following the booster dose.

The contribution of the CD8+T cell response to vaccine efficacy has become increasingly recognized as they have been detected early after vaccination[23] and due to their role in controlling infection[24, 25]. Specifically, it has been established that CD8+T cells expressing cytokines such as IFNγ and TNFα as well as markers involved in activation status and proliferation such as CD38 and Ki67 contribute to limiting disease severity during SARS-CoV-2 infection[24]. Additional studies have identified the expression of CD69 and CD137 on SARS-CoV-2 specific CD8+T cells being associated with less severe disease[25]. This expanded Phase 1 trial demonstrates that immunization with INO-4800 induces SARS-CoV-2 specific CD8+T cells exhibiting these specific characteristics, suggesting the induction of a vaccine-induced cellular response that has potential to protect against severe COVID-19. As VoCs continue to emerge, the generation of cross-reactive activated CD8+T cells with lytic potential is likely to play an important role in preventing severe disease. We have previously demonstrated that vaccination with INO-4800 induces T cells and neutralizing antibodies that are active against the parental SARS-CoV-2 strain as well as the B.1.1.7, B.1.351, and P.1 VoCs[26]. We acknowledge limitations to this trial that include the relatively small study population and the limited number of PBMCs available for testing across more than one assay. This trial was not powered to formally compare immune responses between dose groups or age stratifications. In addition, due to different immune assays and methodologies employed by various groups, it is not possible to directly compare immune responses observed in this trial to those elicited from other vaccine platforms or to determine if the magnitudes observed in this trial are sufficient to confer clinical benefit.

The immune responses observed in the current trial and in our larger Phase 2 trial[15] support advancing the 2.0 mg dose of INO-4800 to a Phase 3 efficacy evaluation. This dose has elicited the highest binding and neutralizing antibody titers, the highest T-cell cytokine production from both CD4+ and CD8+T cells, and the highest expression of markers associated with attenuation of severe COVID-19 on CD8+T cells.

This trial demonstrated that immune responses elicited by a 2-dose primary series of INO-4800 could be boosted by a third dose without safety or tolerability concerns and positions INO-4800 as an important candidate for continued development as a stand-alone SARS-CoV-2 vaccine, as well as for continued examination in combination approaches. The potential ability to administer INO-4800 multiple times, with high tolerability, along with its ease of scalability and thermostability, contribute to its potential value in combatting the COVID-19 pandemic.

## Declaration of Interests

KAK, EB, JA, MG, DA, ACQ, NL, VA, MD, SW, ML, AS, MPM, PP, TM, TRFS, SJR, JL, MD, ASB, JES, JJK, KEB, LMH, JDB, MPM, Jr. report grants from Coalition for Epidemic Preparedness Innovations, during the conduct of the trial; other from Inovio Pharmaceuticals, outside the submitted work. PT, ELR, MP, AJK, FIZ, DF, KL, JE, MA, and DBW report grants from Coalition for Epidemic Preparedness Innovations, during the conduct of the trial. D.B.W. participates in industry collaborations and has received renumeration for individual services. In the interest of disclosure, D.B.W. reports the following paid associations with commercial partners: Pfizer (Advisory Board), Geneos (Advisory, SRA), Advaccine (Advisory) Astrazeneca (Advisory, Speaker), Inovio (BOD, SRA, Stock ownership), Sanofi (Advisory Board), BBI (Advisory Board, SRA). All other authors declare no potential conflicts of interest.

## Funding

This work is funded by the Coalition for Epidemic Preparedness Innovations (CEPI) and Inovio Pharmaceuticals, Inc.

## Data Availability

The preliminary COVID19-001 Phase 1 clinical study datasets are subject to access restriction to protect subject confidentiality as the clinical study is still ongoing.

## Acknowledgments

The investigators and sponsor express their gratitude for the contribution of all the trial participants and the invaluable advice of the independent Data Safety Monitoring Board. Feedback from the peer reviewers is gratefully acknowledged. We also acknowledge the broader support from the various teams within Inovio Pharmaceuticals (Ning Jiang, MD, PhD; Greta Kcomt Del Rio, BS; Alysia Ryan, BS; Dennis Van De Goor, MS; Kelly Morales, BS; Jacob Walton, BS; Srujan Vadlamudi, BS; David Valenta, PhD; EJ Brandreth, MBA; Dan Jordan, BS; Robert J. Juba Jr, MS; Stephen Kemmerrer, BSME, MBA, PE; Timothy Herring, MPH; Susan Duff, BS; and Prasad Kulkarni, PhD, CMPP), the Wistar Institute (Dr. Ziyang Xu and Edgar Tello Ruiz), the National Infections Service, Public Health England (Naomi Coombes, PhD and Mike Elmore, PhD), Hospital of the University of Pennsylvania (Joseph Quinn, RN), and the Alliance for Multispecialty Research, Kansas City, MO and Lexington, KY.

## Supplementary Methods

### Study Design and Enrollment Progression

Participants enrolled at one of three locations in the U.S.: The University of Pennsylvania Clinical Trials Unit in Philadelphia, PA; the Alliance for Multispecialty Research in Kansas City, MO; and the Alliance for Multispecialty Research in Lexington, KY.

### Assessment of SARS-CoV-2 Specific Immune Responses

#### SARS-CoV-2 Spike Enzyme-Linked Immunosorbent Assay (ELISA) description

Costar high bind plates were coated by incubating 1-3 days at 2-8°C with 2.0 µg/mL of recombinant SARS-CoV-2 spike protein trimer (Acro Biosystems; SPN-C52H9) which contains amino acids 16-1213 of the original Wuhan-Hu-1 isolate spike protein (ascension# QHD43416.1). The protein contains six proline substitutions for trimer stabilization and two alanine substitutions to eliminate the furin cleavage site. Coated plates were blocked with Starting Block (Thermo Scientific), followed by the addition of study samples, diluted 1/20 in Starting Block buffer. Plates were incubated for 1 hour, and then washed three times in PBS-tween. 1/1000-diluted anti-human IgG HRP conjugate (BD Pharmingen; 555788) was added, and plates were incubated for an additional hour. Plates were washed three times in PBS-tween, and SureBlue TMB substrate (SeraCare) was added. Assay plates were developed for approximately 9 minutes, and the color development was stopped by the addition of TMB Stop Solution (SeraCare).

SARS-CoV-2 spike antibody concentrations were assigned to study samples by interpolation from a 4-parameter logistic model fit to a dilution curve of SARS-CoV-2 convalescent plasma. The convalescent plasma reference material (Aalto Bio Reagents Ltd) collected from a single convalescent donor >28 days after symptom onset from a PCR-confirmed SARS-CoV-2 test was arbitrarily assigned a concentration of 20,000 Units per mL. This sample was used as a control to standardize IgG responses across participant samples.

#### SARS-CoV-2 Pseudovirus Neutralization Assay

Briefly, heat inactivated study serum samples were serially diluted and incubated for 90 minutes with a SARS-CoV-2-DeltaCT pseudovirus which is based on an HIV/luciferase reporter vector and expresses both SARS-CoV-2 spike protein and luciferase. After incubation, the pseudovirus/serum mixture was added to CHO target cells with stable ACE2 expression. After incubating the pseudovirus, serum, and cells together at 37°C for 72h, a britelite^TM^ Plus luminescence reporter gene assay system (Perkin Elmer) was used to lyse cells and luminescence was measured using a Biotek plate reader.

#### SARS-CoV-2 Spike ELISpot Assay Description

Briefly, 300,000 peripheral mononuclear cells (PBMCs) per well were stimulated overnight in triplicate wells of a pre-coated IFN-γ ELISpot plate (MabTech) using 200 ng per peptide per well (1 µg/mL per peptide final concentration) of a single peptide pool consisting of 15-mer peptides that overlapped by 9 residues and spanned the full-length spike protein. The next day, plates were developed as according to the manufacturer’s instructions (MabTech, Human IFN-g ELISpot Plus). A CTL S6 Micro Analyzer (CTL) with *ImmunoCapture* and *ImmunoSpot* software was used to scan and count spots corresponding to IFN-γ secreting cells. The ELISpot assay was performed in several batches due to the longevity of the study- the week 0 timepoint is run with each post-dose batch in order to have an appropriate baseline control. To most accurately represent the data, the baseline sample is subtracted from the post dose sample run in the same batch. The average of all week 0 values is 8.2 SFU/10^6^ and the median is 3.3 SFU/10^6^.

## Supplementary Data

**Supplementary Table 1:** Geometric Mean Titers (GMT) in the pseudovirus neutralization assay

| Dose Group | Week 0 |  | Week 6 |  |  | Week 8 |  |  | Week 12 |  |  | Week 28 |  |  |
| --- | --- | --- | --- | --- | --- | --- | --- | --- | --- | --- | --- | --- | --- | --- |
|  | N | GMT (95% CI) | N | GMT (95% CI) | P value | N | GMT (95% CI) | P value | N | GMT (95% CI) | P value | N | GMT (95% CI) | P value |
| 0.5 mg | 40 | **3.01**<br>(1.97, 4.60) | 40 | **14.94**<br>(7.90, 28.25) | <0.001 | 40 | **6.00**<br>(3.23, 11.17) | <0.001 | 39 | **3.65**<br>(1.00, 101.0) | 0.018 | 35 | **8.17**<br>(1.00, 106.0) | <0.001 |
| 1.0 mg | 35 | **6.18**<br>(1.00, 235.0) | 35 | **19.07**<br>(8.79, 41.37) | 0.003 | 35 | **14.79**<br>(6.93, 31.55) | 0.005 | 35 | **12.50**<br>(5.75, 27.19) | 0.023 | 21 | **7.00**<br>(2.57, 19.03) | 0.308 |
| 2.0 mg | 36 | **3.50**<br>(2.12, 5.78) | 36 | **54.15**<br>(31.46, 93.21) | <0.001 | 36 | **16.13**<br>(7.39, 35.23) | <0.001 | 36 | **11.37**<br>(5.93, 21.80) | 0.009 | 19 | **11.87**<br>(4.58, 30.76) | 0.036 |

P values compare titers at each study week with those at Week 0

| Dose Group | Pre-boost |  | Post-boost |  |
| --- | --- | --- | --- | --- |
|  | N | GMT (95% CI) | N | GMT (95% CI) |
| 0.5 mg | 28 | **9.6** (4.6, 20.02) | 31 | **58.73** (32.92, 104.75) |
| 1.0 mg | 22 | **7.61** (2.92, 19.84) | 25 | **76.12** (35.18, 164.69) |
| 2.0 mg | 28 | **7.65** (3.75, 15.62) | 30 | **100.0** (51.46, 194.33) |

**Supplementary Table 2:** Geometric Mean Titers (GMT) in ELISA

| Dose Group | Week 0 |  | Week 4 |  |  | Week 6 |  |  | Week 8 |  |  | Week 12 |  |  | Week 28 |  |  |
| --- | --- | --- | --- | --- | --- | --- | --- | --- | --- | --- | --- | --- | --- | --- | --- | --- | --- |
|  | N | GMT (95% CI) | N | GMT (95% CI) | P value | N | GMT (95% CI) | P value | N | GMT (95% CI) | P value | N | GMT (95% CI) | P value | N | GMT (95% CI) | P value |
| 0.5 mg | 40 | **103.55**<br>(78.0, 137.46) | 40 | **173.47**<br>(110.93, 271.25) | 0.035 | 40 | **333.54**<br>(201.16, 553.02) | 0.002 | 40 | **428.45**<br>(279.42, 656.97) | <0.001 | 40 | **330.47**<br>(214.29, 509.66) | 0.001 | 40 | **250.08**<br>(160.91, 388.67) | 0.019 |
| 1.0 mg | 35 | **83.5**<br>(55.04, 126.7) | 35 | **133.89**<br>(76.13, 235.47) | 0.007 | 35 | **337.78**<br>(187.26, 609.28) | <0.001 | 35 | **595.95**<br>(343.01, 1035.40) | <0.001 | 35 | **371.23**<br>(220.08, 626.19) | <0.001 | 25 | **215.26**<br>(113.62, 407.82) | 0.026 |
| 2.0 mg | 36 | **100.69**<br>(70.9, 143.01) | 36 | **236.7**<br>(144.59, 387.48) | 0.004 | 36 | **653.59**<br>(380.01, 1124.13) | <0.001 | 36 | **677.97**<br>(414.95, 1107.72) | <0.001 | 36 | **468.35**<br>(305.26, 718.57) | <0.001 | 19 | **407.15**<br>(239.95, 690.85) | 0.012 |

P values compare titers at each study week with those at Week 0

| Dose Group | Pre-boost |  | Post-boost |  |
| --- | --- | --- | --- | --- |
|  | N | GMT (95% CI) | N | GMT (95% CI) |
| 0.5 mg | 31 | **292.65** (199.21, 429.93) | 31 | **1963.77** (1111.61, 3469.2) |
| 1.0 mg | 29 | **261.71** (156.33, 438.14) | 29 | **3685.49** (1992.72, 6816.2) |
| 2.0 mg | 32 | **285.91** (198.45, 411.91) | 32 | **5953.14** (3616.44, 9799.6) |

**Supplementary Table 3:** Spike-specific SFU/10^6^ PBMCs in the IFN-γ ELISpot assay

| Dose Group | Week 6 |  | Week 8 |  | Week 12 |  | Week 28 |  |
| --- | --- | --- | --- | --- | --- | --- | --- | --- |
|  | N | Median SFU/10<sup>6</sup> PBMCs (95% CI) | N | Median SFU/10<sup>6</sup> PBMCs (95% CI) | N | Median SFU/10<sup>6</sup> PBMCs (95% CI) | N | Median SFU/10<sup>6</sup> PBMCs (95% CI) |
| 0.5 mg | 40 | **19.4**<br>(11.1, 23.3) | 38 | **10.0**<br>(4.4, 25.6) | 36 | **10.6**<br>(6.7, 26.7) | 38 | **1.1**<br>(1.0, 3.3) |
| 1.0 mg | 34 | **7.2**<br>(2.8, 20.0) | 33 | **17.8**<br>(4.4, 27.8) | 32 | **13.3**<br>(6.7, 28.9) | 22 | **6.5**<br>(1.0, 20.6) |
| 2.0 mg | 33 | **43.3**<br>(24.4, 70.0) | 35 | **24.4**<br>(18.9, 57.8) | 32 | **27.8**<br>(11.1, 54.4) | 19 | **19.6**<br>(5.9, 47.8) |

Median of background subtracted magnitudes 95% CI = 95% Non-parametric confidence intervals for the median
| Dose Group | Pre-boost |  | Post-boost |  |
| --- | --- | --- | --- | --- |
|  | N | Median SFU/10<sup>6</sup> PBMCs (95% CI) | N | Median SFU/10<sup>6</sup> PBMCs (95% CI) |
| 0.5 mg | 31 | **6.7** (4.4, 12.2) | 31 | **10.0** (6.7, 18.9) |
| 1.0 mg | 29 | **6.7** (3.3, 12.2) | 28 | **8.3** (3.3, 17.8) |
| 2.0 mg | 33 | **6.7** (3.3, 22.2) | 31 | **16.7** (8.9, 47.8) |

95% CI = 95% Non-parametric confidence intervals for the median

**Supplementary Table 4:** **Summary of treatment-related Adverse Events by Age, Term, and Dose:** Treatment related AEs were reported by A) twelve 18-50 year old participants (20%) reported B) five 51-64 year old participants (16.7%) and C) one ≥65 year old participant (3.3%). All AEs were Grade 1 (mild) in severity with the exception of Grade 2 (moderate) lethargy, abdominal pain, and injection site pruritus. In case of multiple events, a participant is counted only once per system organ class and once per preferred term.

| System Organ Class Preferred Term | 18-50 Years, n (%) |  |  |  |
| --- | --- | --- | --- | --- |
|  | INO-4800 0.5 mg (N=20) | INO-4800 1.0 mg (N=20) | INO-4800 2.0 mg (N=20) | Total (N=60) |
| **At least one related TEAE** | **6 (30.0)** | **4 (20.0)** | **2 (10.0)** | **12 (20.0)** |
| **General disorders and administration site conditions** | **6 (30.0)** | **4 (20.0)** | **1 (5.0)** | **11 (18.3)** |
| Injection site erythema | 1 (5.0) | 3 (15.0) | 0 | 4 (6.7) |
| Injection site pain | 2 (10.0) | 2 (10.0) | 1 (5.0) | 5 (8.3) |
| Injection site pruritus | 3 (15.0) | 0 | 0 | 3 (5.0) |
| Fatigue | 1 (5.0) | 0 | 0 | 1 (1.7) |
| Feeling hot | 1 (5.0) | 0 | 0 | 1 (1.7) |
| Injection site swelling | 0 | 1 (5.0) | 0 | 1 (1.7) |
| **Gastrointestinal disorders** | **0** | **1 (5.0)** | **1 (5.0)** | **2 (3.3)** |
| Abdominal pain upper | 0 | 1 (5.0) | 0 | 1 (1.7) |
| Nausea | 0 | 0 | 1 (5.0) | 1 (1.7) |
| **Musculoskeletal disorders** | **0** | **1 (5.0)** | **0** | **1 (1.7)** |
| Myalgia | 0 | 1 (5.0) | 0 | 1 (1.7) |
| **Nervous system disorders** | **0** | **1 (5.0)** | **0** | **1 (1.7)** |
| Headache | 0 | 1 (5.0) | 0 | 1 (1.7) |
| Lethargy | 0 | 1 (5.0) | 0 | 1 (1.7) |
| **Skin and subcutaneous tissue disorders** | **1 (5.0)** | **0** | **0** | **1 (1.7)** |
| Pruritus | 1 (5.0) | 0 | 0 | 1 (1.7) |
| Rash | 1 (5.0) | 0 | 0 | 1 (1.7) |

B)
| System Organ Class Preferred Term | 51-64 Years, n (%) |  |  |  |
| --- | --- | --- | --- | --- |
|  | INO-4800<br>0.5 mg (N=10) | INO-4800<br>1.0 mg (N=10) | INO-4800<br>2.0 mg (N=10) | Total (N=30) |
| **At least one related TEAE** | **0** | **2 (20.0)** | **3 (30.0)** | **5 (16.7)** |
| **General disorders and administration site conditions** | **0** | **2 (20.0)** | **3 (30.0)** | **5 (16.7)** |
| Injection site pain | 0 | 1 (10.0) | 1 (10.0) | 2 (6.7) |
| Injection site pruritus | 0 | 0 | 1 (10.0) | 1 (3.3) |
| Injection site induration | 0 | 1 (10.0) | 0 | 1 (3.3) |
| Injection site coldness | 0 | 0 | 1 (10.0) | 1 (3.3) |
| Fatigue | 0 | 0 | 1 (10.0) | 1 (3.3) |
| **Respiratory, thoracic, and mediastinal disorders** | **0** | **0** | **1 (10.0)** | **1 (3.3)** |
| Dyspnea | 0 | 0 | 1 (10.0) | 1 (3.3) |

C)
| System Organ Class Preferred Term | ≥65 Years, n (%) |  |  |  |
| --- | --- | --- | --- | --- |
|  | INO-4800<br>0.5 mg (N=10) | INO-4800<br>1.0 mg (N=10) | INO-4800<br>2.0 mg (N=10) | Total (N=30) |
| **At least one related TEAE** | **1 (10.0)** | **0** | **0** | **1 (3.3)** |
| **General disorders and administration site conditions** | **1 (10.0)** | **0** | **0** | **1 (3.3)** |
| Injection site pain | 1 (10.0) | 0 | 0 | 1 (3.3) |

---

**Supplementary Figure 1:**
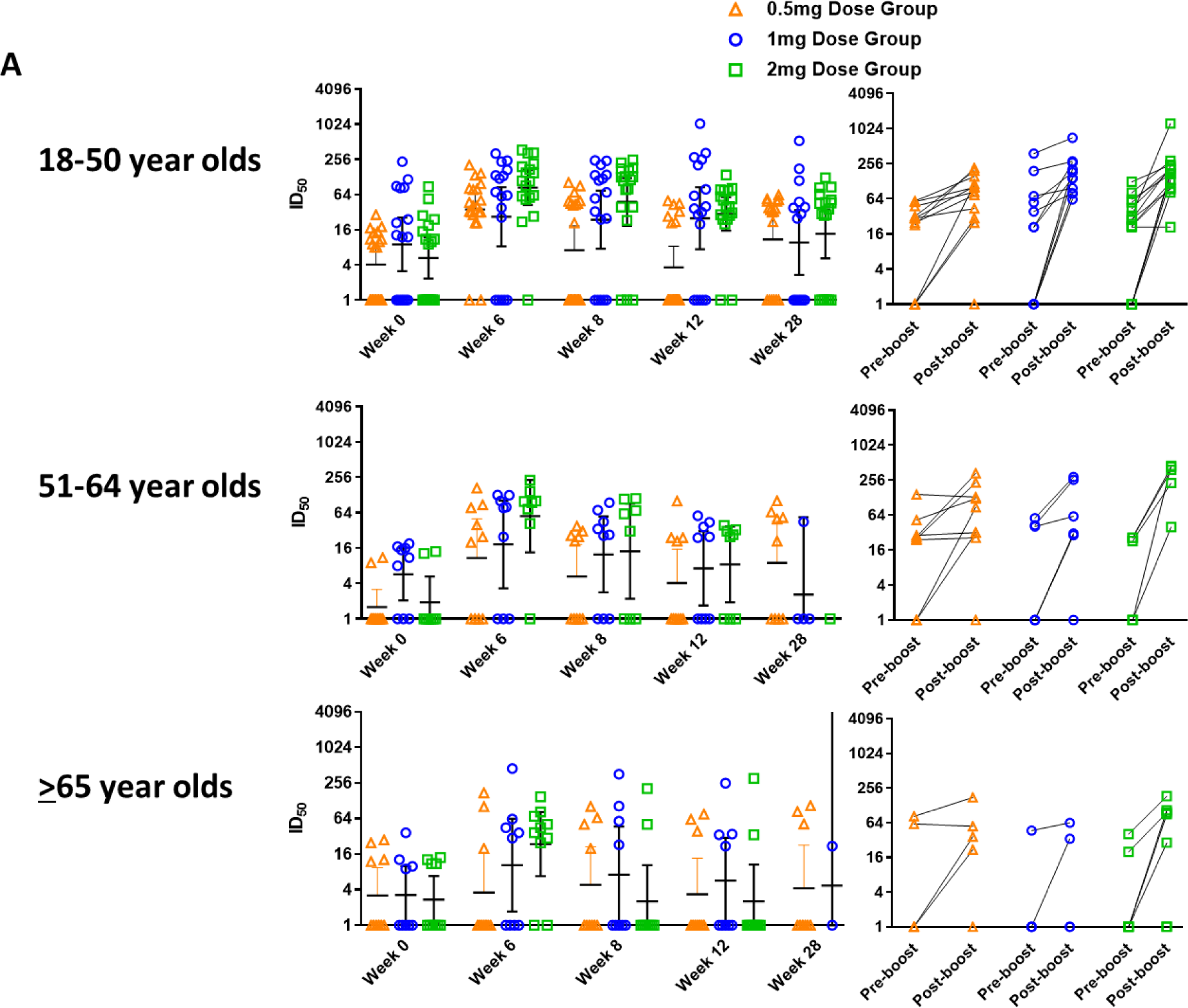
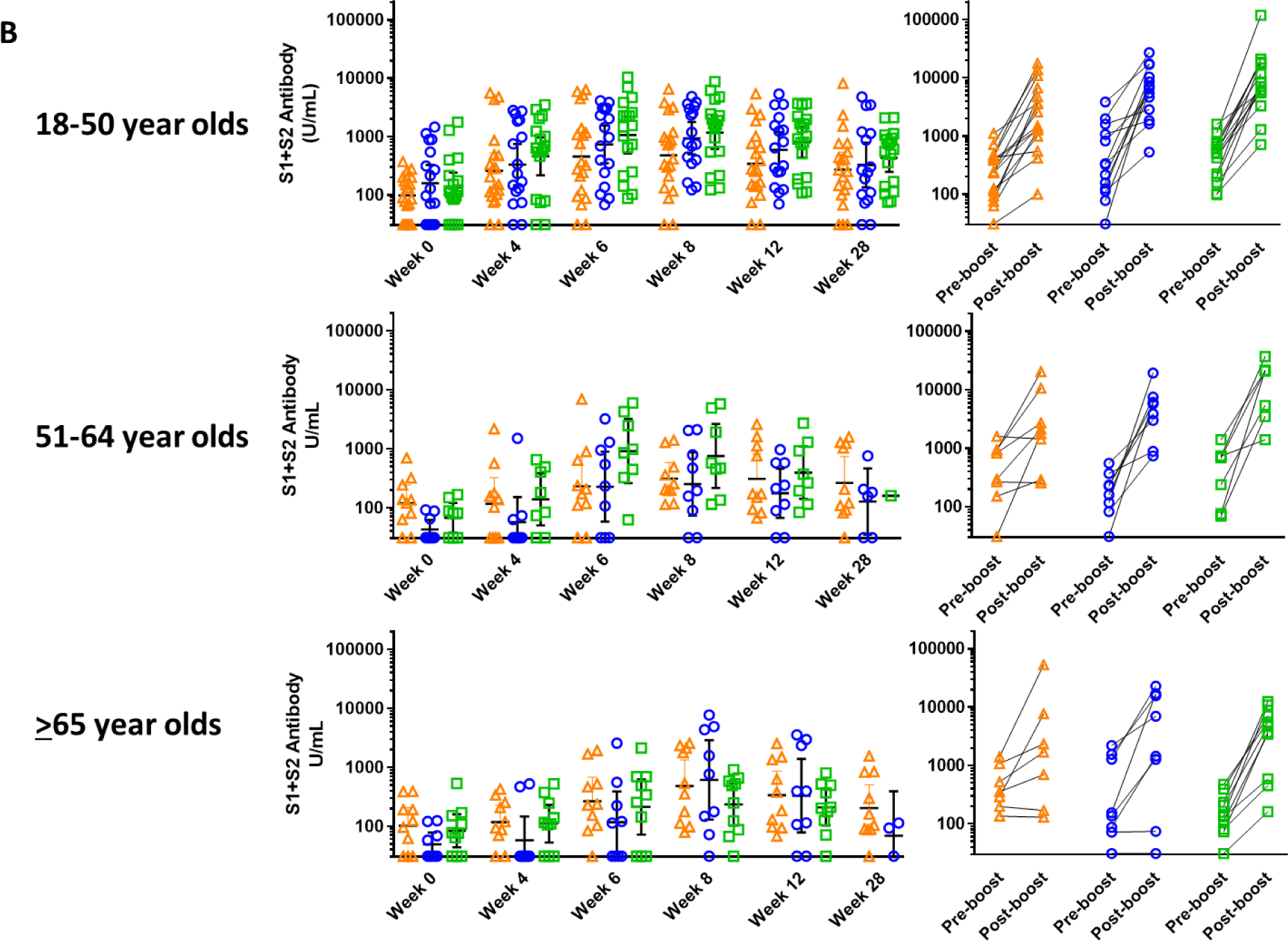
INO-4800 induces antibodies to SARS-CoV-2 across all age groups 18-50, 51-64 and ≥65 year olds A) Functional antibodies were assessed using a pseudovirus neutralization assay. The inhibition dilution where 50% neutralization occurs (ID_50_) is plotted. The left panel includes n=40 participants in the 0.5 mg dose group (n=20 18-50 year olds, n=10 51-64 year olds and n=10 ≥65 year olds), n=35 participants in the 1.0 mg dose group (n=17 18-50 year olds, n=9 51-64 year olds and n=9 ≥65 year olds) and n=36 participants in the 2.0 mg dose group (n=18 18-50 year olds, n=8 51-64 year olds and n=10 ≥65 year olds). The right panel includes only participants with paired data available n=26 (n=12 18-50 year olds, n=9 51-64 year olds and n=5 ≥65 year olds), n=21 (n=11 18-50 year olds, n=6, 51-64 year olds and n=4 ≥65 year olds) and n=27 (n=15 18-50 year olds, n=5 51-64 year olds and n=7 ≥65 year olds) participants in the 0.5 mg, 1.0 mg, and 2.0 mg dose groups, respectively. B) Binding antibody concentrations to the Spike trimer were measured using ELISA. The left panel includes n=40 participants in the 0.5 mg dose group (n=20 18-50 year olds, n=10 51-64 year olds and n=10 ≥65 year olds), n=35 participants in the 1.0 mg dose group (n=17 18-50 year olds, n=9 51-64 year olds and n=9 ≥65 year olds) and n=36 participants in the 2.0 mg dose group (n=18 18-50 year olds, n=8 51-64 year olds and n=10 ≥65 year olds). The right panel includes only participants with paired data available n=31 (n=16 18-50 year olds, n=8 51-64 year olds and n=7 ≥65 year olds), n=29 (n=13 18-50 year olds, n=8, 51-64 year olds and n=8 ≥65 year olds) and n=32 (n=15 18-50 year olds, n=7 51-64 year olds and n=10 ≥65 year olds) participants in the 0.5 mg, 1.0 mg, and 2.0 mg dose groups, respectively. Open symbols represent individual participants, the horizontal line represents the GMT and the whiskers extend to the 95%CI values. Paired t test was used to assess significance versus baseline. The dose groups are represented by orange triangles (0.5 mg), blue circles (1.0 mg) and green squares (2.0 mg).

**Supplementary Figure 2:**
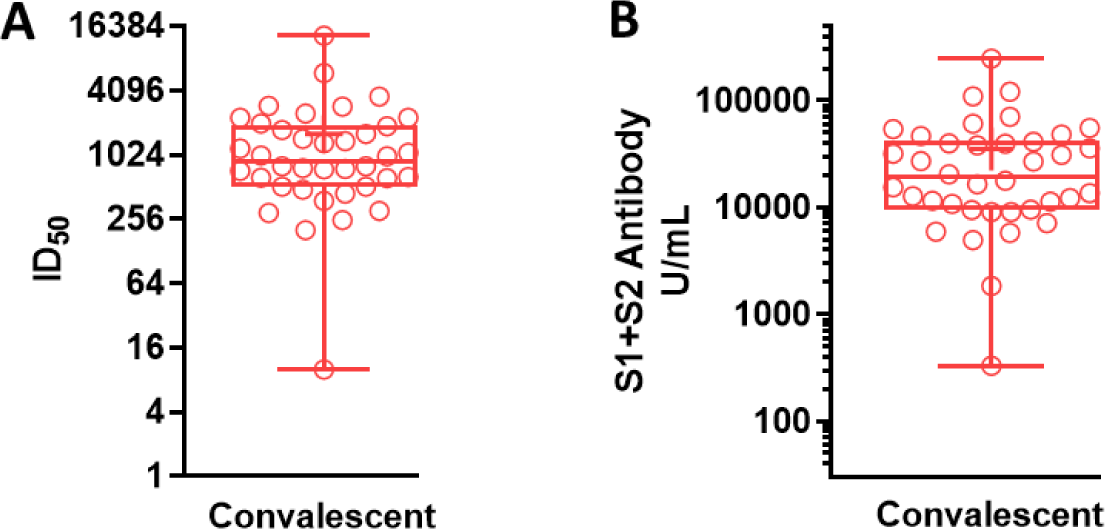
Antibody responses in SARS-CoV-2 convalescent donors. Plasma from convalescent donors (n=38) was obtained. A) Functional antibodies were assessed using a pseudovirus neutralization assay. The inhibition dilution where 50% neutralization occurs (ID_50_) is plotted. B) Binding antibody concentrations to the Spike trimer were measured using ELISA.

**Supplementary Figure 3:**
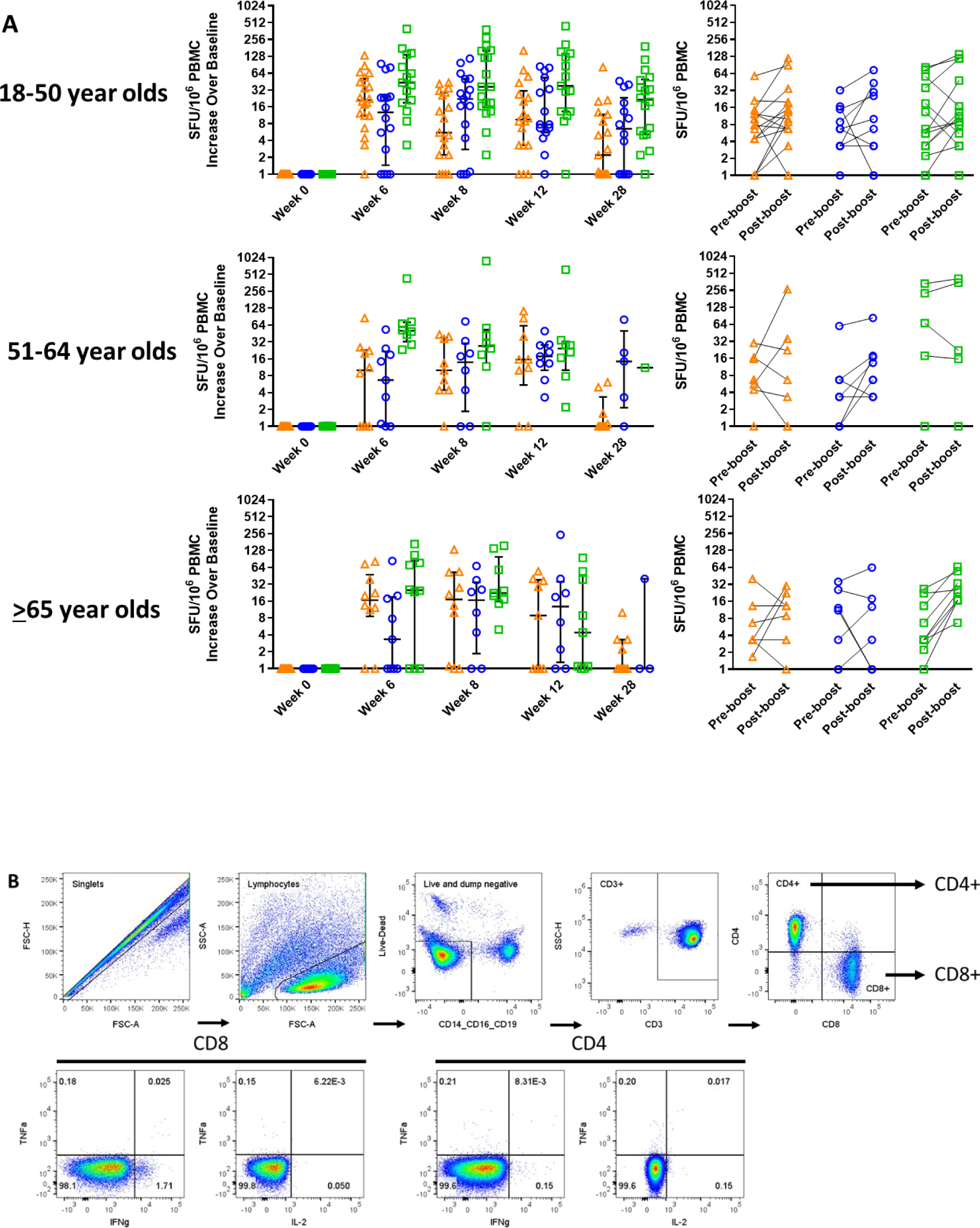
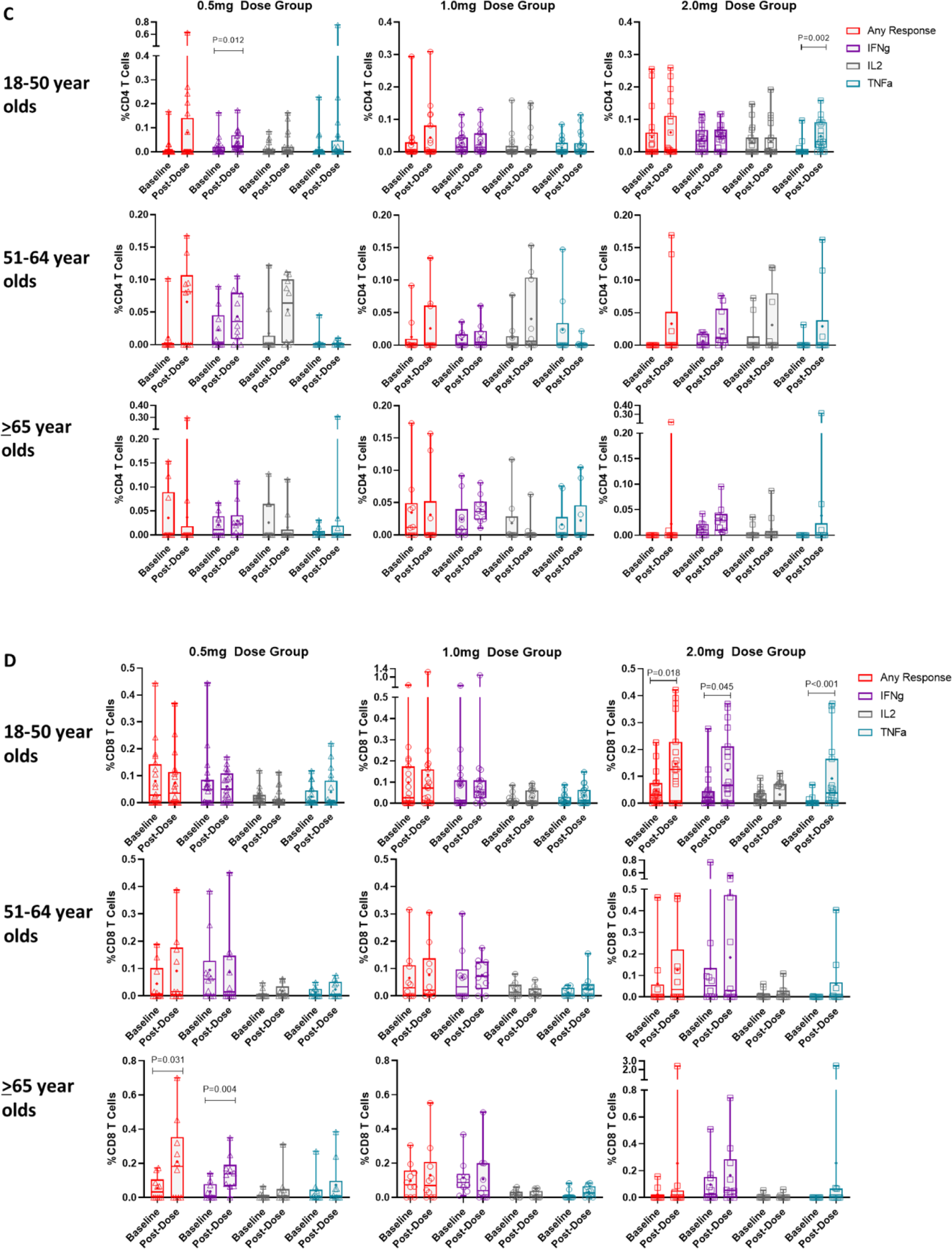
INO-4800 induces cellular responses to SARS-CoV-2 Spike across all age groups (18-50, 51-64, and ≥65 years). A) Longitudinal increases in spike antigen specific spot forming units per 10^6^ PBMCs over baseline in the IFN-g ELISpot are plotted. The left panel includes n=40 participants in the 0.5 mg dose group (n=20 18-50 year olds, n=10 51-64 year olds and n=10 ≥65 year olds), n=35 participants in the 1.0 mg dose group (n=17 18-50 year olds, n=9 51-64 year olds and n=9 ≥65 year olds) and n=36 participants in the 2.0 mg dose group (n=18 18-50 year olds, n=8 51-64 year olds and n=10 ≥65 year olds). The right panel includes only participants with paired data available n=34 (n=19 18-50 year olds, n=8 51-64 year olds and n=7 ≥65 year olds), n=24 (n=10 18-50 year olds, n=7, 51-64 year olds and n=7 ≥65 year olds) and n=28 (n=14 18-50 year olds, n=5 51-64 year olds and n=9 ≥65 year olds) participants in the 0.5 mg, 1.0 mg, and 2.0 mg dose groups, respectively. B) Example gating strategy for flow cytometry assays. C-D) Intracellular cytokine staining for IFN-g (purple) IL-2 (gray), TNF-a (blue) or any of the three cytokines (red) are plotted from samples collected at baseline or post-dose 2. The graphs include n=40 participants in the 0.5 mg dose group (n=20 18-50 year olds, n=10 51-64 year olds and n=10 ≥65 year olds), n=35 participants in the 1.0 mg dose group (n=19 18-50 year olds, n=10 51-64 year olds and n=10 ≥65 year olds) and n=36 participants in the 2.0 mg dose group (n=19 18-50 year olds, n=10 51-64 year olds and n=10 ≥65 year olds). Open symbols represent individual participants, the box extends from the 25^th^ to the 75^th^ percentile, line inside the box is the median, and the whiskers extend from the minimum to maximum values. The mean is denoted with a “+” sign. Wilcoxon signed-rank was used to assess significance versus baseline. The dose groups are represented by triangles (0.5 mg), circles (1.0 mg) and squares (2.0 mg).

## Notes

### Clinical Trial

NCT04336410

### Author Declarations

The trial was approved by the institutional review board of each clinical site (The University of Pennsylvania Clinical Trials Unit in Philadelphia, PA; the Alliance for Multispecialty Research in Kansas City, MO; and the Alliance for Multispecialty Research in Lexington, KY.) and all participants provided written informed consent prior to enrollment. The trial was conducted under current Good Clinical Practices (GCP).

### Summary of Updates

Revisions made for clarification. Supplemental files updated.

